# Donor HLA class 1 evolutionary divergence and late allograft rejection after liver transplantation in children: An emulated target trial

**DOI:** 10.1101/2024.09.13.24313304

**Authors:** Jérôme Dumortier, Sarah Hamada, Emma Wischlen, Céline Mandier, Noémie Laverdure, Olivier Boillot, Ilias Kounis, Vincent Allain, Valérie Hervieu, Sophie Collardeau-Frachon, Valérie Dubois, Cyrille Feray

**Affiliations:** Hospices Civils de Lyon, Hôpital Edouard Herriot, Fédération des Spécialités Digestives, Lyon; Université Claude Bernard Lyon 1, Lyon; Etablissement Français du Sang, Décines; Hospices Civils de Lyon, Hôpital Femme-Mère-Enfants, Service d’Hépato-gastroentérologie et Nutrition, Bron; AP-HP, Hôpital Paul Brousse, Centre Hépato-Biliaire, INSERM, Unité 1193, Hepatinov, and Université Paris-Saclay, Villejuif; Hôpital Saint-Louis, Assistance Publique-Hôpitaux de Paris (AP-HP), Université de Paris, Laboratoire d’Immunologie et Histocompatibilité, Paris; Hospices Civils de Lyon, Groupe hospitalier Est, Service d’Anatomie pathologique, Bron, France

**Author notes:** Correspondance: Jérôme Dumortier, Pavillon L, Hôpital Edouard Herriot, 69437 Lyon Cedex 03, France. Financial support: There was no funding source for this study. **Primary Funding Source:** None.

**Keywords:** liver transplantation, rejection, HLA evolutionary divergence, anti-HLA antibodies

## Abstract

HLA evolutionary divergence (HED), a continuous metric quantifying the differences between each amino acid of two homologous HLA alleles, reflects the importance of the immunopeptidome presented to T lymphocytes. It has been associated with rejection after liver transplantation. This retrospective cohort study aimed to analyze the potential effect of donor or recipient HED on liver transplant rejection in a new series of patients transplanted during childhood and followed in adulthood. The study included 120 children who had been transplanted between 1991 and 2010 and were followed by routine biopsies and histological evaluations with a median of 14.1 years post-LT. Liver biopsies were performed routinely 1, 5, 10 and 20 years after transplantation and in the event of liver dysfunction. HED was calculated using the physicochemical Grantham distance for donor and recipient class I (HLA-A, -B, -C) and class II (HLA-DRB1, -DQB1) alleles. The influence of HED on rejection was analyzed using IPW and target trial emulation using the g method. Based on the IPW score, donor HED class I was correlated with the occurrence of late (>90 days) rejection (HR, 1.19, 95% CI: 1.01-1.40) independently of HLA mismatches, donor age and initial induction. This emulated target trial confirmed that donor HED class I has a causal effect on liver graft rejection and this relationship was observed long-term.

## Introduction

HLA plays an essential role in allograft rejection, and the importance of HLA compatibility between donors and recipients has been well established (1). Regarding HLA, a more recent concept relies on the difference between the two alleles (2). The more divergent the two HLA alleles at a physicochemical level (Grantham’distance (3), the greater the size of the immunopeptidome. This is an extension of the idea of heterozygote advantage (4). HLA evolutionary divergence (HED) has emerged as a strong determinant of survival in patients with cancer who have received immune checkpoint inhibitors (5)(6) and in those with leukemia who have undergone allogeneic hematopoietic stem cell transplants (7).

In the context of organ transplantation, HED may increase the diversity of graft-derived immunopeptidomes targeted by recipient T-cells. In an adult population and in a smaller series of transplanted children from Necker Hospital, we previously reported that regardless of HLA compatibility, HED defined from donor class I HLA was positively associated with rejection (8).

The target trial approach is a structural method that emulates an RCT (9) based on observational data, and enables the definition of a causal relationship between exposure and a given outcome. In the present study, we performed an emulated target trial to analyze the influence of HED on the occurrence of late histological signs of rejection in patients transplanted during childhood and adulthood.

## Methods

### Study Population

Characteristics of the patients and donors are shown in Table 1.

### Procedure

Liver biopsies were performed in the event of abnormal liver test results and systematically at 1, 5, 10, 15 and 20 years after LT. The liver specimens were paraffin-embedded and stained with hematein-eosin-safran and picrosirius. The diagnosis of acute or chronic rejection was made according to the Banff Classification of Allograft Pathology (10) (11), while acute cellular rejection was classified using the Banff Rejection Activity Index. Chronic rejection was based on bile duct dystrophy, ductopenia, and hepatic venule fibrosis. Late chronic rejection was based on a ductopenia rate of 50% or more. The histological diagnoses were coded prospectively. The diagnosis of antibody-mediated rejection was based on the positive C4d staining of portal veins and portal capillaries and positive DSA.

### Biomarkers

Donor-specific antibodies were available pre- and post-LT using Luminex Single Antigen Flow Beads assays (LSA class I and class II; Lifecodes, Immucor, Norcross, GA, US). Mean fluorescence intensity (MFI) was measured on a LABscan IS 200, and all specificities were evaluated using the company’s defined threshold, i.e. MFI ≥1500 and AD-BCR ≥5 to define positivity (AD-BCR is the ratio of adjusted MFI to the quantity of coated antigen per bead).

HLA typing for HLA-A,-B,-C,-DRB1 and -DQB1 was available for 120 donor/recipient couples at a low-resolution level (one field) (the HLA typing of donors and recipients was performed using Luminex PCR-SSO reverse, One Lambda, Canoga Park, CA, US) and then extrapolated to a high-resolution level (two fields) using a tool called HaploSFHI (12). HLA matching was calculated for each recipient as the number of HLA-A, HLA-B, HLA-C, HLA-DRB1 and HLA-DQB1 identities with the donor. Sequence divergence (at the amino acid level) between HLA alleles was computed for all possible combinations of allele pairs among the alleles found in both cohorts for HLA-A, HLA-B, HLA-C, HLA-DRB1, and HLA-DQB1 loci. The respective protein sequences of the peptide-binding groove (exons 2 and 3 for HLA class I and exon 2 for HLA class II) were extracted from the international ImMunoGeneTics/HLA database (13). The calculation of HED between aligned allele pairs of a given locus was based on the Grantham distance metric (3). For each recipient and each donor, the mean class I HED and mean class II HED were calculated as the mean of HED defined at the HLA-A, HLA-B and HLA-C and HLA-DRB1 and HLA-DQB1 loci, respectively, assuming that each locus contributes equally to the presentation of peptides. By definition, there was null divergence in the case of homozygosity. HED values were calculated in 2023 so they did not influence any diagnostic or therapeutic decisions.

### Statistical Analysis

#### IPW approach

To estimate the marginal effect of HED on the occurrence of rejection, we used the inverse probability weighting approach precisely described in (8) (14) based on covariate balancing generalized propensity scores (CBGPS) (15). First, we defined a set of demographic and clinical variables based on their clinical relevance. We followed the recommendations of Fong and colleagues (15) to add the squares of the continuous variables in the CBGPS and use a Box-Cox transformation of continuous exposure (16). Once fitted, the individual stabilized weightings were obtained from the CBGPS (14). Second, we fitted a weighted, cause-specific, proportional hazard Cox model with the exposure (i.e. HED) as the only explanatory variable (17). E-values were computed to assess the exchangeability assumption— in other words, no unmeasured confounding (18).

**Emulated target trial used G computation**, a causal inference technique which is possible on binary exposure (19) to estimate the average treatment (i.e., marginal causal) effect on the entire population (ATE). The ATE is the average effect, at the population level, of moving an entire population from untreated to treated. In this study, the treatment was to receive or not receive a liver from a high HED donor and the outcome was rejection-free survival. The survival area plot (20) directly depicts the survival probability over time and as a function of a continuous covariate which is HED. This plot utilizes g-computation based on a suitable time-to-event model to obtain the relevant estimates. By using g-computation, these estimates can be adjusted for confounding and permit a causal interpretation under the standard causal identifiability assumptions.

All analyses were performed using R, version 3.6.0 (R Foundation) with the “survival,” “CBPS,” “ipw,” “splines,” “EValue,” “RISCA”, “forestplot” and “contsurvplot” packages.

### Ethics

This study was approved by the local Ethics Committee and was performed in accordance with the principles of the Declaration of Helsinki. As it concerned a retrospective cohort without therapeutic intervention, no informed consent of the patients was required under French law (Jardé Law).

## Results

The flow chart (Figure 1) describes patient selection. We only included patients who had undergone at least one liver biopsy, and we excluded premature deaths. The characteristics of the study population are shown in Table 1. The median number of biopsies was five [IQR: 3-7].

**Table 1.** Characteristics of the study population (n=120)

| **Characteristic** | **Value** |
| --- | --- |
| **Recipient** |  |
| Median age (IQR), y | 4.68 (1.10-6.20) |
| Female, *n* (%) | 68 (56.7%) |
| UNOS status, *n* (%) |  |
| 1 | 16 (13.3%) |
| 2A | 9 (7.5%) |
| 2B | 52 (43.3%) |
| 3 | 43 (35.8%) |
| Child-Pugh score, *n* (%) |  |
| A | 19 (15.8%) |
| B | 61 (50.8%) |
| C | 40 (33.3%) |
| Principal indication, *n* (%) |  |
| Biliary atresia | 63 (52.5%) |
| Genetic liver disease <sup>a</sup> | 27 (22.5%) |
| Metabolic disorder <sup>b</sup> | 11 (9.2%) |
| Fulminant hepatitis | 8 (6.7%) |
| Tumor | 1 (0.8%) |
| Autoimmune | 3 (2.5%) |
| Other indications <sup>c</sup> | 7 (5.8%) |
| Combined kidney graft, *n* (%) | 12 (10.0%) |
| **Donor/graft, n** |  |
| Median age (IQR), y | 21 (12-31) |
| Female, *n* (%) | 48 (40.0%) |
| Living donor, *n* (%) | 38 (31.7%) |
| Full liver, *n* (%) | 40 (33.3%) |
| Left lobe, *n* (%) | 71 (59.2%) |
| Left liver, *n* (%) | 5 (4.2%) |
| Right liver, *n* (%) | 4 (3.3%) |
| Median cold ischemic time (IQR), *min* | 425 (151-632) |
| Number of HLA identities, *n* (%) |  |
| 0–2 | 58/118* (49.2%) |
| 3–8 | 60/118* (50.8%) |
| **Liver graft biopsy** |  |
| Median number (IQR) | 6 (4.25-8.75) |
| Abbreviations: HBV = hepatitis B virus; HCV = hepatitis C virus; IQR = interquartile range<br>UNOS = United Network for Organ Sharing.<br>a. Progressive familial intrahepatic cholestasis (n=8), α-1-antitrypsin deficiency (n=4),<br>Alagille syndrome (n=6), Primary sclerosing cholangitis (n=5), Isolated congenital hepatic<br>fibrosis (n=2), Wilson's disease (n=7)<br>b. Hyperoxalosis (n=10), Crigler Najjar syndrom (n=1), Methylmalonic Acidemia (n=1)<br>c. Budd-Chiari syndrome (n=1), Cystic Fibrosis (n=2), C virus-related cirrhosis (n=1),<br>NASH (n=1), Unknown (n=4)<br>*HLA class II was not available for 2 patients |  |

**Figure 1.**
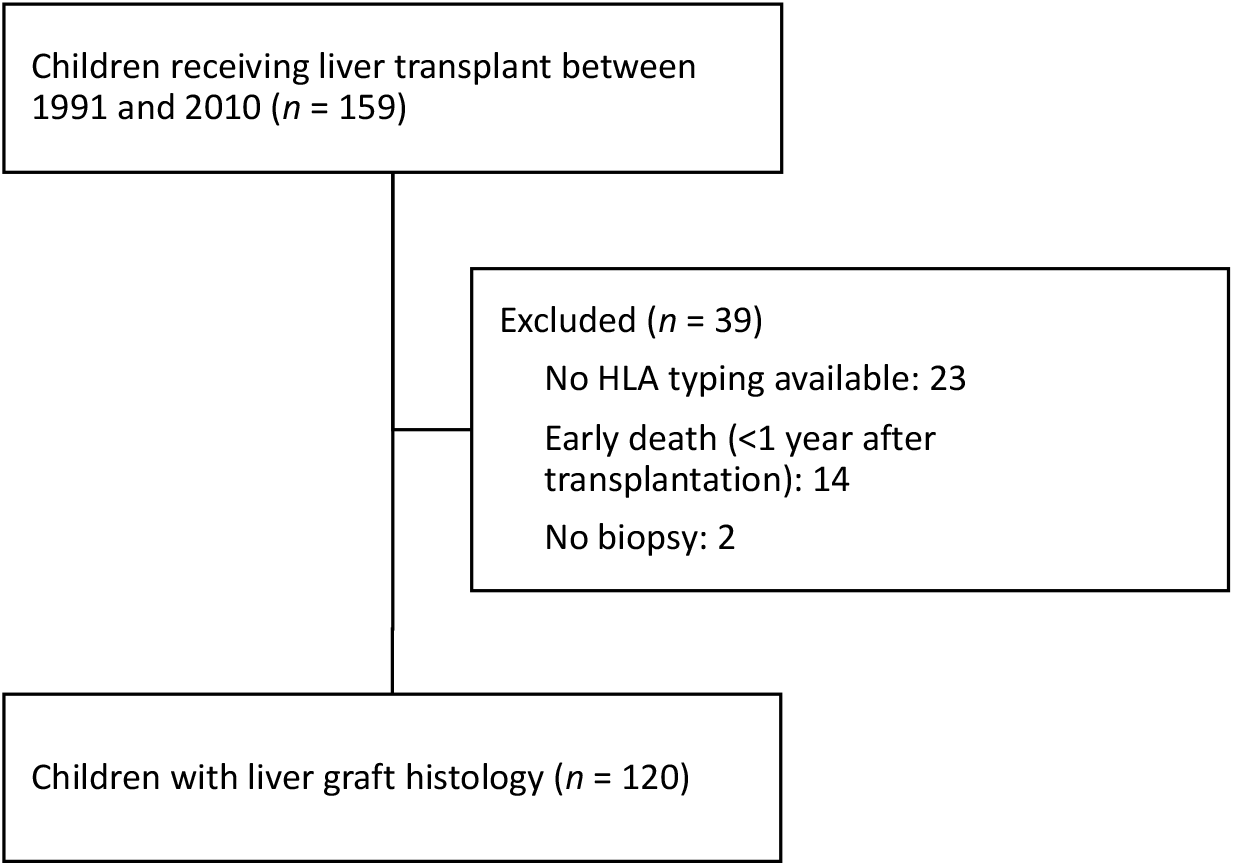
Flow chart of patients included in the study.

During histological follow-up, 88/120 patients (73%) presented with histological signs of rejection. Fifty-one of them displayed early signs of histological rejection (≤90 days), so that once these patients had been censored, only 37 could be evaluated for rejection beyond 90 days post-LT. Because all patients with early biopsies had signs of rejection, we analyzed the cumulative incidence of rejection on biopsies harvested beyond 90 days. Rejection lesions were encountered after 90 days in 64 patients, were purely acute in 41 patients, and chronic in 23, 14 of whom had also signs of acute rejection. In four cases of CR, the rejection was of a humoral type.

Through the univariate log-rank analysis of rejection-free survival, we selected six covariates (Figure 2) associated with the occurrence of rejection: HLA matching, initial induction, recipient gender, donor age, recipient age and cold ischemia time (more or less than 385 min). Among all 17 covariates, none was correlated or associated with any measured donor or recipient HED. These six covariates were used subsequently for IPW and for the emulated target trial.

**Figure 2.**
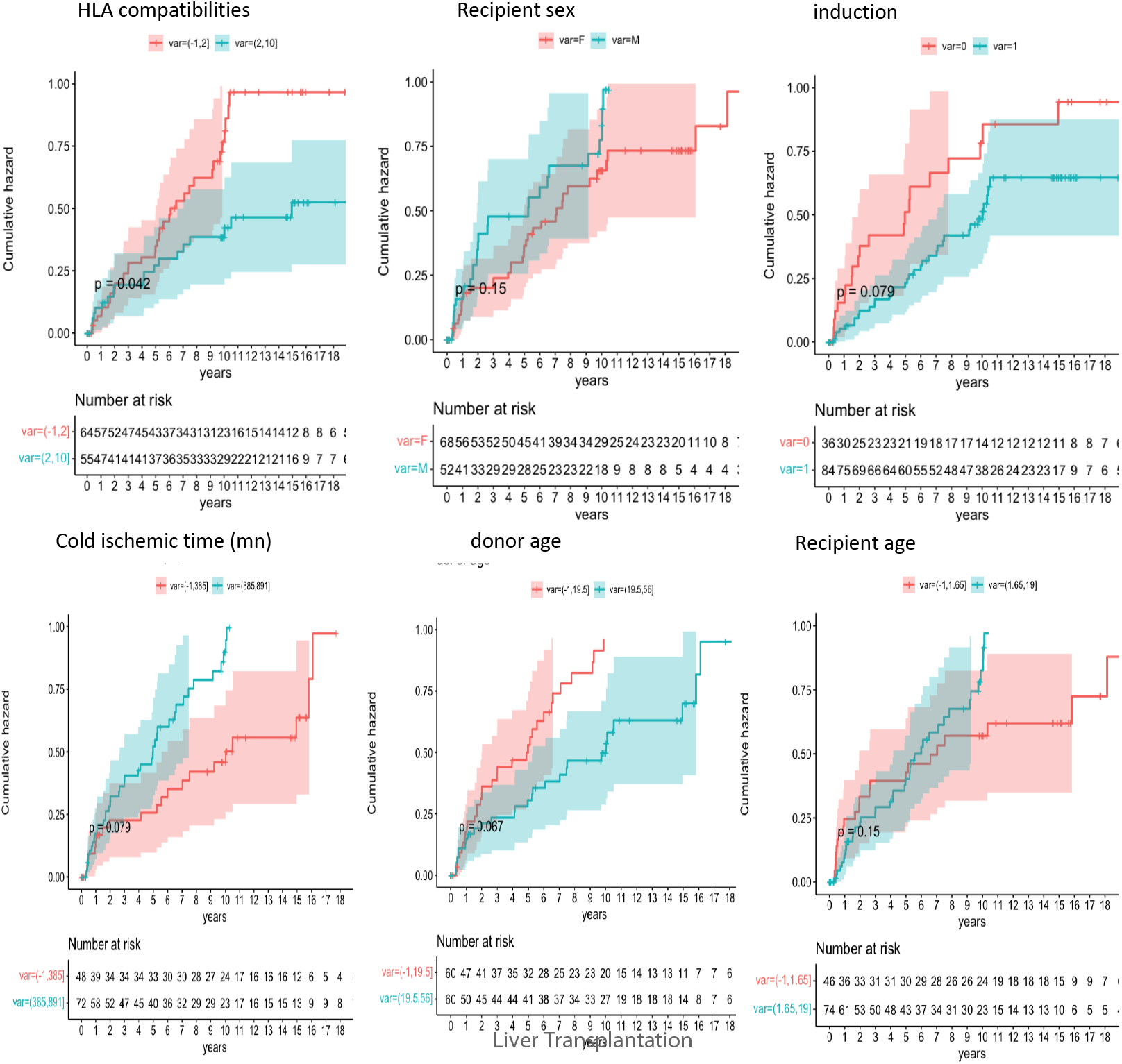
Impact of HLA incompatibilities, recipient gender, initial induction, cold ischemia time, donor age and recipient age on the risk of rejection over time.

HED values were considered as continuous for the IPW approach. As HED were not normally distributed, a Box-Cox transformation was performed. For the propensity score, we checked the balance of covariates and distribution of stabilized weightings as being correct. As shown in Figure 3, donor HED class I was positively associated with rejection while donor HED class II and recipient HED were not.

**Figure 3.**
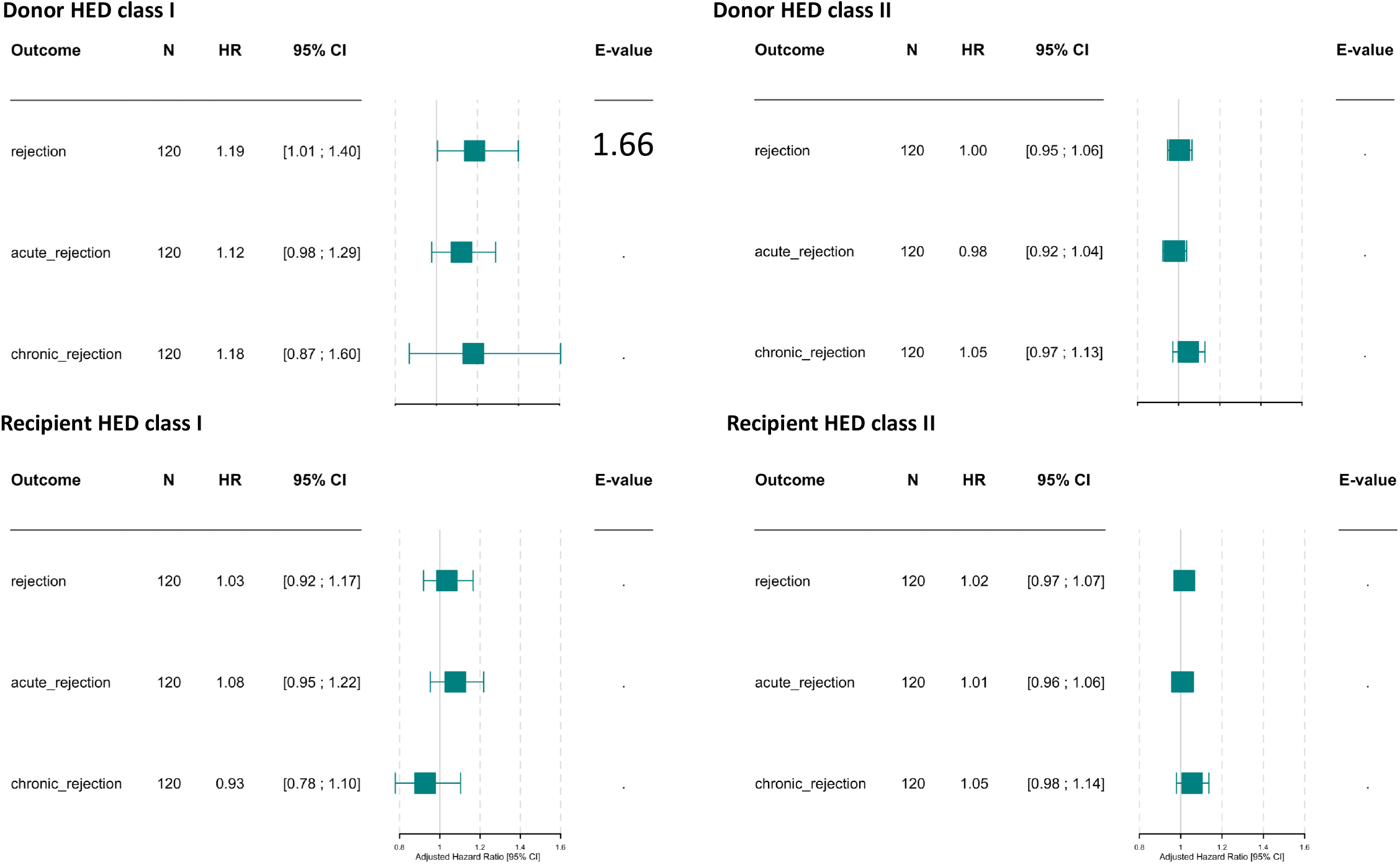
IPW approach applied to the effect of donor and recipient class I and class II HED on rejection, acute rejection and chronic rejection. The results show adjusted hazard ratio (HR) values with a 95% confidence interval (CI) and E-values. Donor class I HED was associated with late rejection but not with either acute or chronic rejection. All other HEDs were not associated with rejection..

For the emulated target trial, the exposure needed to be binary as it was simulating a randomized trial. Since donor HED class I (as a continuous variable) was positively associated with rejection, we tested different thresholds for HEDs. Using the contsurvplot package we were able to illustrate the effect of donor HED class I on the occurrence of rejection (Figure 4A). Figure 4B shows the Odds ratio for each emulated trial. The risk of rejection was increased at values of 7 or higher.

**Figure 4.**
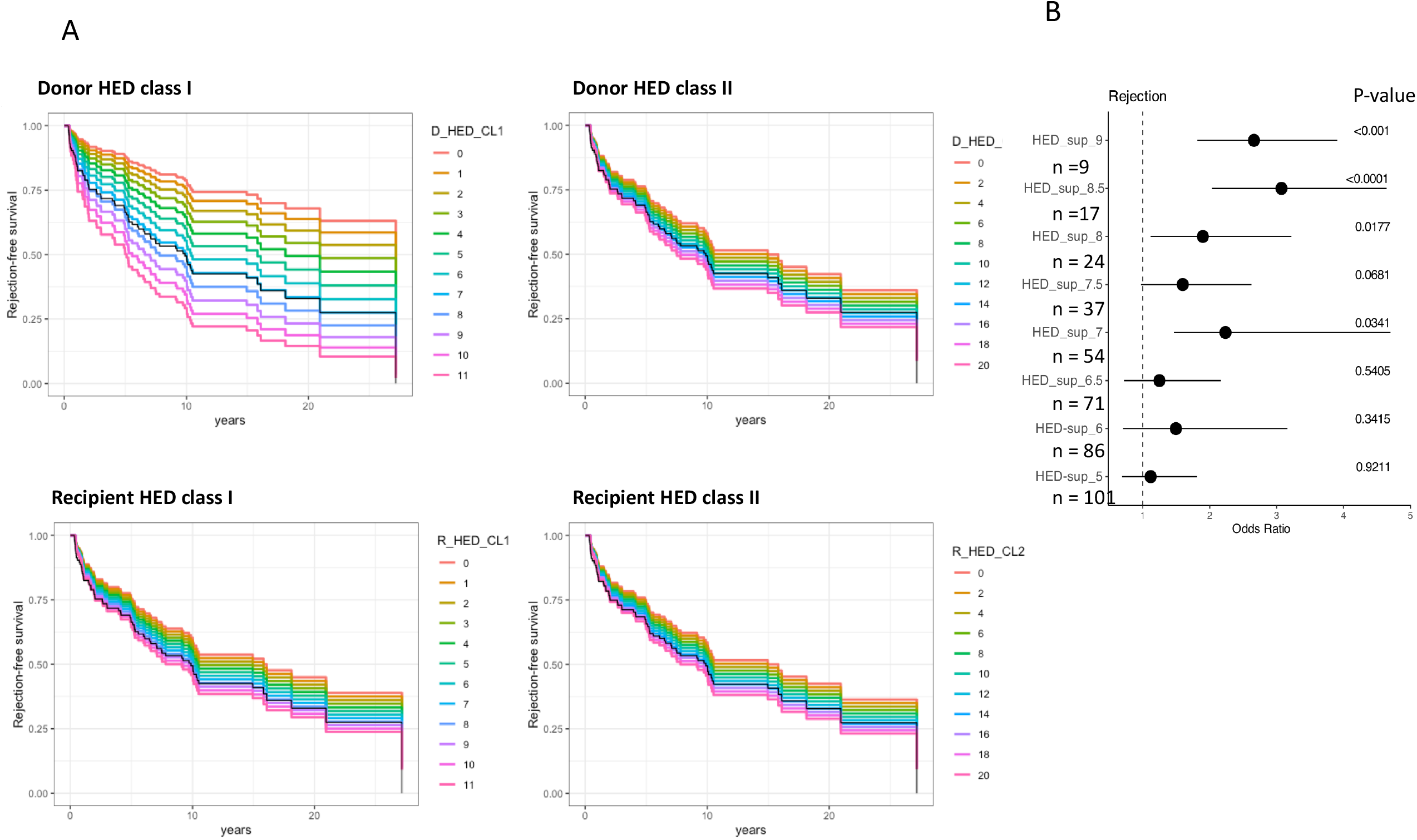
Emulated Target Trial using g computation applied to the risk of rejection with different values of donor or recipient HED class I or II.

Finally, the presence of DSA was not related to any HEDs (as a continuous or categorial variable) and was not related to rejection.

## Discussion

This study confirms the findings of our previous study (8) showing that donor HED class I is a predictor of rejection after liver transplantation. However, in the present series, the patients had all been transplanted as children and the majority were studied in adulthood after a longer follow-up, thus suggesting that the effect of HED on rejection persists over time. The methodology was also enhanced. As in the first study (8), we used the IPW approach to study HED as a continuous variable. But furthermore, we performed an emulated target trial that simulated a therapeutic trial (receiving or not receiving a graft with HED class I >7) that met the standard causal identifiability assumptions (14). For this reason, a causal relationship between an HED class I donor and late rejection could be confirmed in the context of LT.

The relationship between donor HED and rejection was observed when early biopsies (e.g. <90 days) were excluded. Early (<90 days) rejection lesions after liver transplantation are common in children (21)(22). In the previously published pediatric series at Necker Hospital, early biopsies were infrequent, while they were frequent in the present study. This is due to the decline of early biopsies in pediatric centers, the Necker series being more recent than the present Lyon series. By excluding early biopsies from the current series, a direct comparison with the results at Necker Hospital could be made, and according to our findings, it appeared that an HED class I donor plays a role in late rejection.

As in the previous paper, we did not evidence any roles for recipient HED or class II HLA. This is surprising because the alloantigen presentation by a recipient’s or donor’s immune cells is crucial for rejection. However, no genotyping of HLA-DQA1 or -DP was available in this series, nor in the previous one.

One limitation of this study was that we were unable to distinguish between acute and chronic rejection, probably because of the small sample size and the low incidence of pure chronic rejection. In addition, we did not find a relationship between HED and DSA, despite the availability of the latter. This had also been the case in the Necker series. One possible reason for this might be the long study period, with heterogeneity affecting the tests used to detect DSA after TH (measurement bias). The other reason could be that the HLA-DQA1 locus was not explored and might possibly be related to the immune response. We recently published an article concerning the crucial influence of this locus on the response to HBV and SARS-CoV-2 vaccines (23).

There have been few recent advances regarding the immunology of liver transplantation. The demonstration that donor HED class I affects rejection suggests that this metric should be considered in clinical practice. In patients with a high risk of rejection, the selection of a “low HED donor” might be more feasible than selection based on HLA compatibilities. It is worth noting that donor HED class I is not correlated with HLA donor/recipient compatibility and that the effect of donor HED class I was independent of HLA compatibility. The proposed metric could also guide the monitoring of biopsies and aid in selecting patients who may either be suitable for weaning from immunosuppression (24) or require significant long-term immunosuppression.

In conclusion, the present study confirms the major role of donor HED class I in liver allograft rejection. This opens a broad horizon for future investigations in organ transplantation, focused in particular on patients with a higher risk of rejection-associated graft loss, such as kidney or lung transplants. Further studies involving larger cohorts could, for instance, investigate acute and chronic rejection separately, the occurrence of DSA, or immune tolerance.

## Supporting information

graphicalAbstract

## Data Availability

All data produced in the present study are available upon reasonable request to the authors

## Abbreviations

CBGPS: covariate balancing generalized propensity scores
CNI: Calcineurin-Inhibitors
CSA: Cyclosporine
DSA: Donor Specific Antibody
HED: HLA Evolutionary Divergence
HLA: Human Leucocyte Antigen
IPW: Inverse Probability Weighting
MMF: Mycophenolate Mofetil
TAC: Tacrolimus

