## Supplementary material for "Donor HLA class 1 evolutionary divergence and late allograft rejection after liver transplantation in children: An emulated target trial": graphicalAbstract

### Slide 1
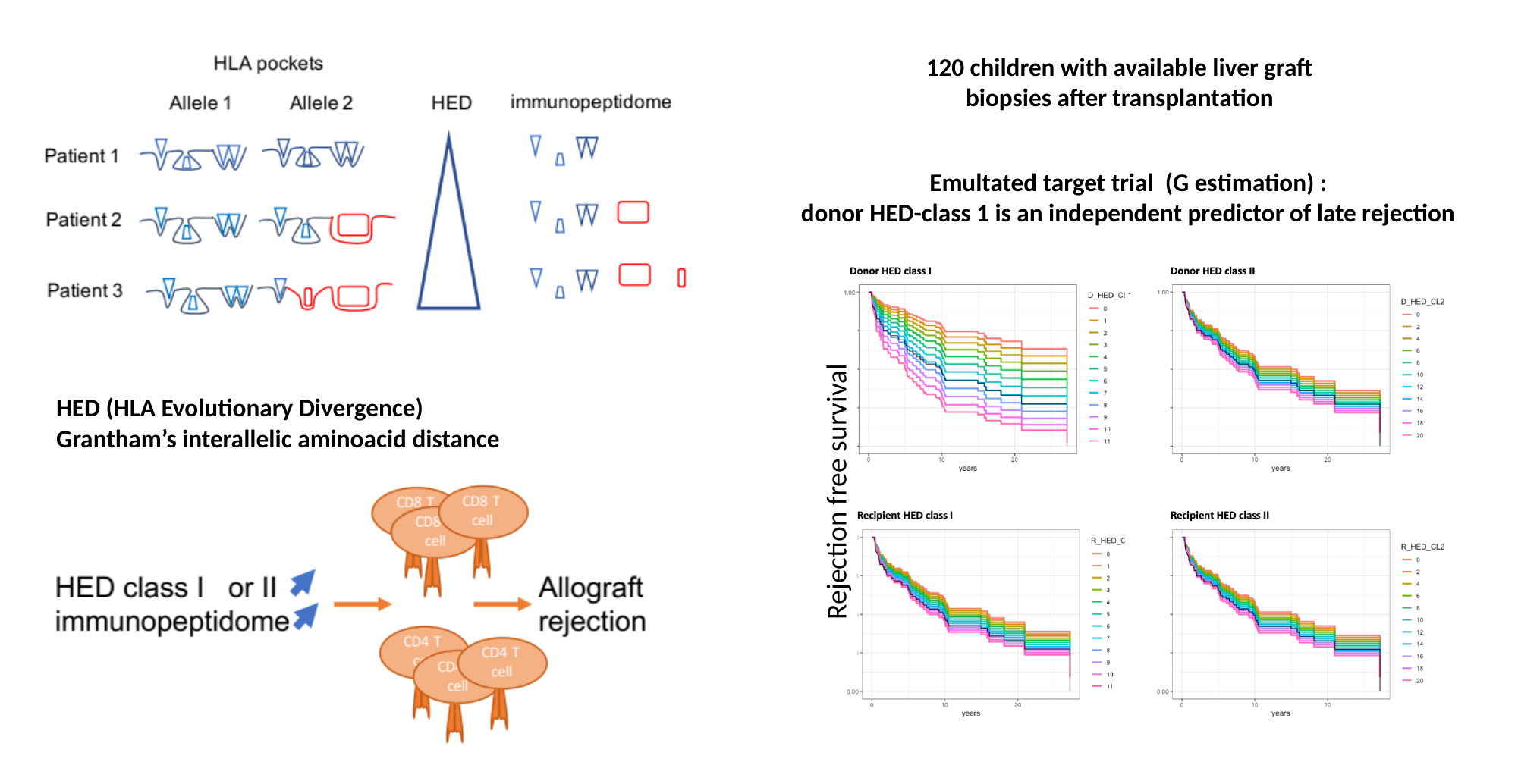

120 children with available liver graft biopsies after transplantation
Emultated target trial (G estimation) :
donor HED-class 1 is an independent predictor of late rejection
HED (HLA Evolutionary Divergence)
Grantham’s interallelic aminoacid distance
Rejection free survival
Rejection free survival
